# Shared Decision Making for Patients Hospitalized with Acute Myocardial Infarction. A Randomized Trial

**DOI:** 10.1101/2022.02.17.22271131

**Authors:** Megan E. Branda, Marleen Kunneman, Alejandra I. Meza-Contreras, Nilay D. Shah, Erik P. Hess, Annie LeBlanc, Jane A. Linderbaum, Danika M. Nelson, Margaret R. Mc Donah, Carrie Sanvick, Holly K. Van Houten, Megan Coylewright, Sara R. Dick, Henry H. Ting, Victor M. Montori

## Abstract

**Objective:** Adherence to guideline-recommended medications after acute myocardial infarction (AMI) is suboptimal. Patient fidelity to treatment regimens may be related to their knowledge of the risk of death following AMI, the pros and cons of medications, and to their involvement in treatment decisions. Shared decision making may improve both patients’ knowledge and involvement in treatment decisions.

**Methods:** In a pilot trial, patients hospitalized with AMI were randomized to the use of the *AMI Choice* conversation tool or to usual care. *AMI Choice* includes a pictogram of the patient’s estimated risk of mortality at 6 months with and without guideline-recommended medications, i.e., aspirin, statins, beta-blockers, and angiotensin-converting enzyme inhibitors. Primary outcomes were patient knowledge and conflict with the decision made assessed via post-encounter surveys. Secondary outcomes were patient involvement in the decision-making process (observer-based OPTION12 scale) and 6-month medication adherence.

**Results:** Patient knowledge of the expected survival benefit from taking medications was significantly higher (62% vs. 16%, *p*<0.0001) in the *AMI Choice* group (n=53) compared to the usual care group (n=53). Both groups reported similarly low levels of conflict with the decision to start the medications (13 (SD 24.2) vs. 16 (SD 22) out of 100; *p*=.16). The extent to which clinicians in the *AMI Choice* group involved their patients in the decision-making process was high (OPTION12 score 53 out of 100, SD 12). Medication adherence at 6-months was relatively high in both groups and not different between groups.

**Conclusions:** The *AMI Choice* conversation tool improved patients’ knowledge of their estimated risk of short-term mortality after an AMI and the pros and cons of treatments to reduce this risk. The effect on patient fidelity to recommended medications of using this SDM tool and of SDM in general should be tested in larger trials enrolling patients at high risk for nonadherence.

**Trial registration number:** NCT00888537

## INTRODUCTION

Over 1 million Americans suffer an acute myocardial infarction (AMI) annually and survivors remain at increased risk of adverse outcomes including death, reinfarction, and readmission.^1^ At the time of the present study, at hospital discharge, guidelines recommended that patients be prescribed a “medication bundle” comprised of aspirin, statin, a beta-blocker, and an angiotensin-converting enzyme inhibitor.^2,3^ However, up to 50% of patients discontinue these medications within weeks to months.^4-6^

Poor fidelity to treatments is multifactorial and attributable to both structural factors (e.g., access to and cost of medications) and volitional factors (e.g., treatment is inconsistent with patients’ knowledge and preferences). Recent studies to address structural factors by eliminating out-of-pocket expenses for medications resulted in marginal improvement in adherence rates.^7^ Volitional factors may play an important role in adherence and include reasons such as lack of patient knowledge about their risk for adverse outcomes after hospital discharge, the magnitude of this risk, and the expected benefit from taking medications. We hypothesized that patients hospitalized for AMI, if educated regarding their personalized risk for mortality and the potential benefit of taking medications with proven efficacy, would have greater knowledge, and greater patient involvement in the decision-making process. This involvement makes possible the work that clinicians and patients do together to co-create treatment plans. This work, which takes place in conversations between clinicians and patients, is known as shared decision making (SDM).^8^

To test this hypothesis, we developed the *AMI Choice* conversation tool and evaluated its efficacy in a pilot randomized trial to improve (a) patient knowledge of personalized risk of mortality after AMI and benefit from taking the “medication bundle” and (b) patient involvement in SDM.

## METHODS

### Study design, Setting and Participants

We conducted a single center, prospective randomized pilot trial to compare usual care with and without use of the *AMI Choice* conversation tool at St. Mary’s Hospital, Mayo Clinic, Rochester, MN. Eligible patients were adults who were admitted for AMI and who were expected to discharge to home and could give written informed consent and use the conversation tool (i.e., read English, have no uncorrectable visual or hearing impairment, and have no substantial cognitive impairment as judged by the treating clinicians). The Mayo Clinic Institutional Review Board approved all study procedures.

A computer-generated allocation sequence randomly assigned patients in a concealed fashion using a secure study website, 1:1 to either intervention (*AMI Choice* conversation tool) or to usual care.^9^ Patients were told that the trial was designed to assess and improve communication about medications, but were kept blind about its goal of comparing the efficacy of *AMI Choice* to usual care. Blinding of other study personnel was not implemented.

### Intervention

The *AMI Choice* conversation tool was developed using participatory action research, a method our team has used to create other effective conversation tools for use during face-to-face clinical encounters.^10,11^ This conversation tool displayed a patient’s estimated 6-month risk of mortality after AMI calculated using the GRACE risk score^12^ and the expected reduction in the risk of death associated with the use of the “medication bundle” prescribed by the clinical team. The design features of this conversation tool (**Figure 1**) include: (a) 1000-person pictorial chart to display risk and risk reduction; (b) a tailored estimate of (i) the 6-month mortality based on the GRACE score, and (ii) the absolute risk reduction conferred by medications for which the patient was eligible and prescribed^13^; and (c) a description of the potential harms and costs associated with prescribed medications. Nurse practitioners or registered clinical nurses caring for the patient used the conversation tool together with the patient on the day prior to or on the day of discharge from the hospital. Prior to conducting the study, study coordinators prepared the personalized *AMI Choice* conversation tool and trained the nurses and nurse practitioners to use the conversation tool and to follow a standard script. A clinical cardiologist led a 1-hour training session with participating nurses and an opportunity for role-play of the conversation tool usage with a mock patient. Additionally, study coordinators offered point-of-care coaching lasting 5-10 minutes immediately before use of the conversation tool. Patients in the usual care group discussed medications with their clinical team in the typical manner without any additional support or training. All enrolled patients also received generic pamphlets about the indications, side effects, and costs of each medication.

**Figure 1.**
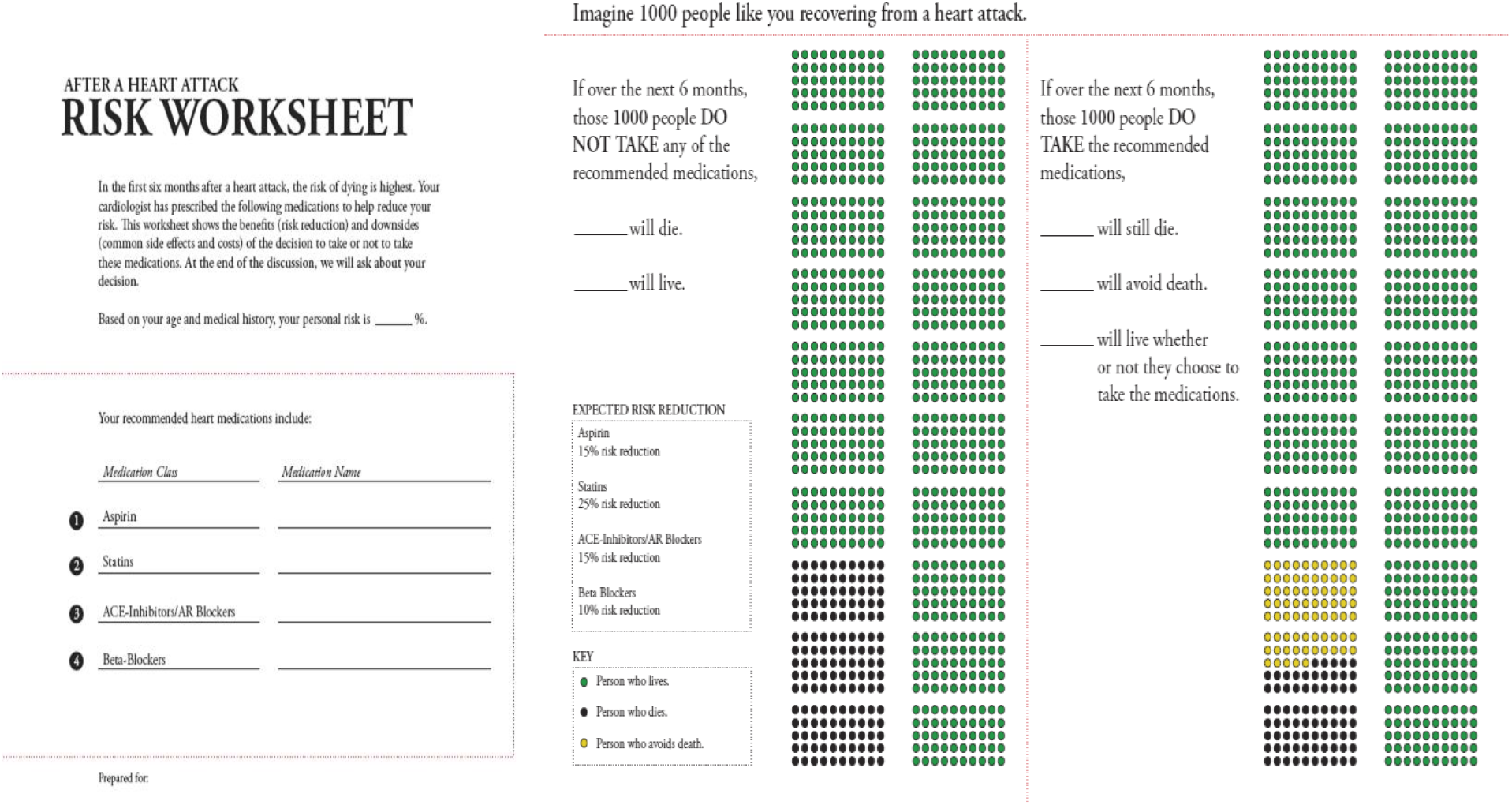
*AMI Choice* Conversation tool for a patient with a 6-month mortality risk of 15%

### Data Collection and Outcomes

Data collection included immediate post-encounter surveys to assess patient knowledge, comfort with the decision made, and delivery of the *AMI Choice* tool or usual care. We conducted follow-up telephone interviews to assess medication initiation and adherence at 5 weeks and 6 months post hospital discharge. We also reviewed pharmacy records of prescribed medications from 3-months before admission to 6 months after discharge. We collected sociodemographic information about each patient and assessed their health literacy using the Rapid Estimate of Adult Literacy in Medicine (REALM).^14^

Patient knowledge was measured by an immediate post-encounter survey, including the following 3 questions: (i) How many people like you, recovering from a heart attack, do you think will die in the next 6 months if they do not take any of the recommended heart medicines? (ii) How many people like you, recovering from a heart attack, do you think will be alive in 6 months if they do not take any of the recommended heart medicines? (iii) How many people like you, recovering from a heart attack, do you think will be alive in 6 months because they took heart medicines? Answer format invited entering a number to complete the sentence “out of 1000 people like you”.

Patient satisfaction with the information presented — its amount, clarity, helpfulness, and the degree with which patients would like to use a similar tool for other decisions and would recommend the tool to others facing the same decision – was captured using 5 questions with a 7-point Likert scale answer with higher scores indicating agreement or strong agreement. Patient conflict with the decision made was assessed by a survey using 8 of the 16 items of the Decisional Conflict Scale.^15^ The Decisional Conflict Scale assesses the level of conflict that an individual experiences when facing a difficult decision that involves competing options with potential benefits and risks.^15^ This modified version assessed patients’ confidence in their knowledge of the information and support received, and the resulting decisional efficacy and satisfaction, with higher scores indicating greater conflict with the decision made.^16^

Video recordings of encounters allocated to the intervention group were used to assess the extent to which clinicians involved their patients in the decision-making process when using the *AMI Choice* tool. We coded these recordings using the OPTION12 scale,^17^ an observer-based, 12-items scale to quantify clinicians’ behavior to involve patients in decision making. Scores are reported on 0-100 scale, with higher scores indicating more behaviors to involve patients. Our group has extended the use of this tool to video recordings with excellent inter-rater reliability (intraclass correlation coefficient > 0.7).^18-20^ As this was a limited-scope pilot trial, only encounters from the conversation tool group were recorded and assessed in this fashion. Normative data from a meta-analysis of prior conversation tool studies suggest that usual care clinicians achieve a median degree of patient involvement of 23 out of 100 in the OPTION12 scale.^21^

We evaluated medication adherence by calculating the percentage of days covered (PDC), defined as the number of days a patient had a supply of each medication divided by the number of days of eligibility for that medication. We also calculated the proportion of patients who were adherent (defined as PDC covered ≥80%) to each and to all three drug classes (angiotensin-converting enzyme inhibitors, beta-blockers, and statins) throughout the follow-up period.

Adherence to aspirin was assessed via self-report using a standard question,^22^ as use of this medicine cannot be ascertained through pharmacy profile. Different medications within a drug class were considered interchangeable.

### Statistical Methods

In prior SDM trials, we have detected meaningful differences in the primary outcome measures (patient knowledge and decisional comfort) with sample sizes of 100 patients and we deemed this sufficient to produce evidence to determine whether to pursue a larger trial to assess efficacy for the outcome of adherence.^23-25^ We did not conduct formal power calculations. Patient knowledge was compared for each of the three questions by comparing the responses to their calculated risk and benefits. We compared knowledge between the two groups by comparing how many patients responded, “Don’t know”, how many were able to report their actual risks and benefits, and how many were able to report their risk and benefits within absolute 5% of the calculated risk and benefits. Patient satisfaction with the information presented and decisional comfort were compared using the mean responses on a 7-point Likert scale and by comparing the mean scores on the Decisional Conflict Scale between the two groups. Hypotheses were tested using Wilcoxon rank-sum tests to compare medians and chi-square or Fisher exact tests to compare frequencies.

All analyses were based on 2-sided tests at a significance level of 0.05. Consistent with the intention-to-treat principle,^26^ we analyzed all patients in the groups to which they were randomized. All analyses were conducted using SAS v 9.2 (SAS Institute, Inc, Cary, NC).

## RESULTS

We enrolled and randomly allocated 107 patients into this pilot study from March 2009 to June 2010. (**Figure 2**); one patient allocated to the usual care group declined further research procedures shortly after enrollment, leaving 106 patients to conduct the trial. Baseline characteristics were well balanced between the intervention (*SDM Choice* conversation tool) and usual care groups (**Table 1**), except for age, current smoking status, and creatinine levels. In this trial, based on the video recorded encounters, use of the conversation tool required an average of 5.5 minutes.

**Figure 2.**
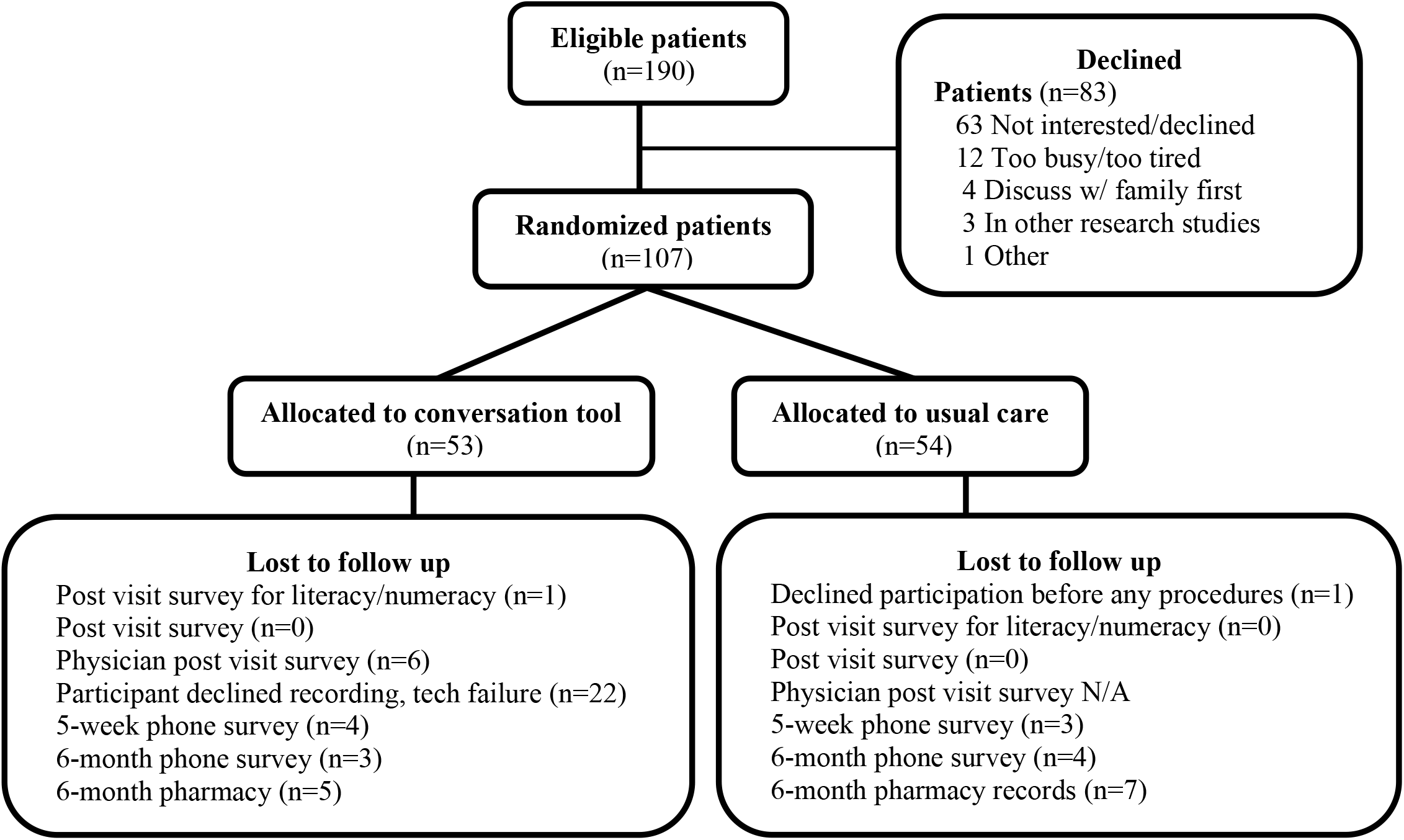
Flow of participants through the trial

**Table 1.** Patient baseline characteristics

|  | Conversation tool<br>(n=53) | Usual Care<br>(n=53) |
| --- | --- | --- |
| Male, n (%) | 37 (70%) | 37 (70%) |
| Age, mean (SD) | 66 (9.0) | 62 (10.5) |
| Married, n (%) | 36 (68%) | 42 (79%) |
| Annual income in US\$, n (%) | | |
| < 20,000 | 4 (7.6) | 5 (9.4) |
| 20-60,000 | 32 (60) | 24 (45) |
| > 60,000 | 13 (25) | 21 (40) |
| Missing | 4 (7.6) | 3 (5.7) |
| Education level, n (%) |  |  |
| High School Diploma or less | 19 (36) | 19 (36) |
| Some College | 12 (23) | 19 (36) |
| 4 Years College/Postgraduate | 15 (28) | 9 (17) |
| Missing | 7 (13) | 6 (11) |
| Smoking status, n (%) |  |  |
| Current smoker | 9 (17) | 20 (38) |
| Past smoker | 27 (51) | 14 (26) |
| Never | 17 (32) | 17 (32) |
| Missing | 0 (0) | 2 (3.8) |
| Comorbidities, n (%) |  |  |
| Congestive heart failure | 4 (7.6) | 3 (5.7) |
| Diabetes | 8 (15) | 13 (25) |
| Percutaneous coronary intervention during index hospitalization, n (%) | 43 (81) | 45 (85) |
| Creatinine (mg/dL), mean (SD) | 1.12 (0.78) | 0.92 (0.33) |
| GRACE Score 6-month mortality, n (%) |  |  |
| Low (< 5%) | 11 (21) | 10 (19) |
| Moderate (5-10%) | 39 (74) | 40 (76) |
| High (> 10%) | 3 (5.7) | 3 (5.7) |
| REALM Score, mean (SD)* | 63.5 (5.6) | 63.8 (3.8) |
| Number of medications at hospital discharge, mean (SD) | 7.5 (2.5) | 7.7 (2.6) |

\* REALM scores were missing from 5 usual care patients and 7 intervention patients.

### Knowledge transfer

Patients using *SDM Choice* were more likely to provide correct responses to survey questions assessing their risk of mortality (50% vs. 2%; *p*<0.001) and their risk reduction from the “medication bundle” (62% vs. 16%; *p*<.001) when compared to those receiving usual care (**Table 2**).

**Table 2.** Patient knowledge by trial group

| | Group | Response exactly correct | *Fisher's exact test* | Response correct ± 5% | *Fisher's exact test* |
| --- | --- | --- | --- | --- | --- |
| Knowledge of likelihood of death in 6 months with no heart medicines. | Usual Care | 2% | $p<.0001$ | 2% | $p<.0001$ |
|  | Conversation tool | 50% |  | 50% |  |
| Knowledge of likelihood of being alive in 6 months with no heart medicines | Usual Care | 2% | $p<.0001$ | 6% | $p<.0001$ |
|  | Conversation tool | 46% |  | 54% |  |
| Knowledge of likelihood of being saved by heart medicines in 6 months. | Usual Care | 0% | $p=.0013$ | 16% | $p<.0001$ |
|  | Conversation tool | 19% |  | 62% |  |

### Participant satisfaction, patient conflict, and patient involvement in treatment decisions

There was no significant difference in patient satisfaction with the way information was shared or in patient-reported decisional conflict between the two groups (**Table 3**). Across all satisfaction questions, 80-100% of participating clinicians (n=5) agreed or strongly agreed with statements in favor of the intervention and its ongoing use, while disagreeing about the likelihood that other colleagues and the institution would support its ongoing use. The extent to which clinicians in the *AMI Choice* group (n=32) involved their patients in the decision-making process based on analysis of the videorecorded encounters was very high (OPTION12 mean score 53, SD 12).

**Table 3.** Conversation tool acceptability and effect on decisional conflict, trust, and knowledge

|  | **Conversation tool**<br>**(n=53)** | **Usual care**<br>**(n=53)** | **Difference**<br>**(95% CI)** |  |
| --- | --- | --- | --- | --- |
| **Satisfaction with knowledge transfer, n (%)** |  |  |  | ***p* value<sup>1</sup>** |
| Information amount | 51 (96) | 45 (85) |  | 0.08 |
| Information clear | 37 (70) | 33 (62) |  | 0.41 |
| Information helpful | 33 (62) | 35 (66) |  | 0.69 |
| Would want same for other decisions | 40 (78)** | 43 (83)* |  | 0.58 |
| Recommend to others | 42 (81)* | 38 (73)* |  | 0.35 |
| **Decisional Conflict Scale, mean (SD)** | 13.0 (24.2) | 16.0 (22.0) | - 3.0 (-11.9, 5.9) | 0.51<sup>2</sup> |

\*, responses missing from 1 participant; \*\* responses missing from 2 participants; 1, Wilcoxon Rank Sum Test; 2, Kruskal-Wallis Test

### Medication initiation and adherence

**Table 4** presents the initiation and continuation information for prescribed medications. Based on the baseline patient survey, 52 of the 53 patients in the conversation tool group decided to start some or all the prescribed medications (n=1 missing), while 50 of the 53 patients in the usual care group decided to start some or all of the prescribed medications (n=1 missing; n=1 did not start any of the medications; n=1 did not believe they were offered the medications).

**Table 4.**
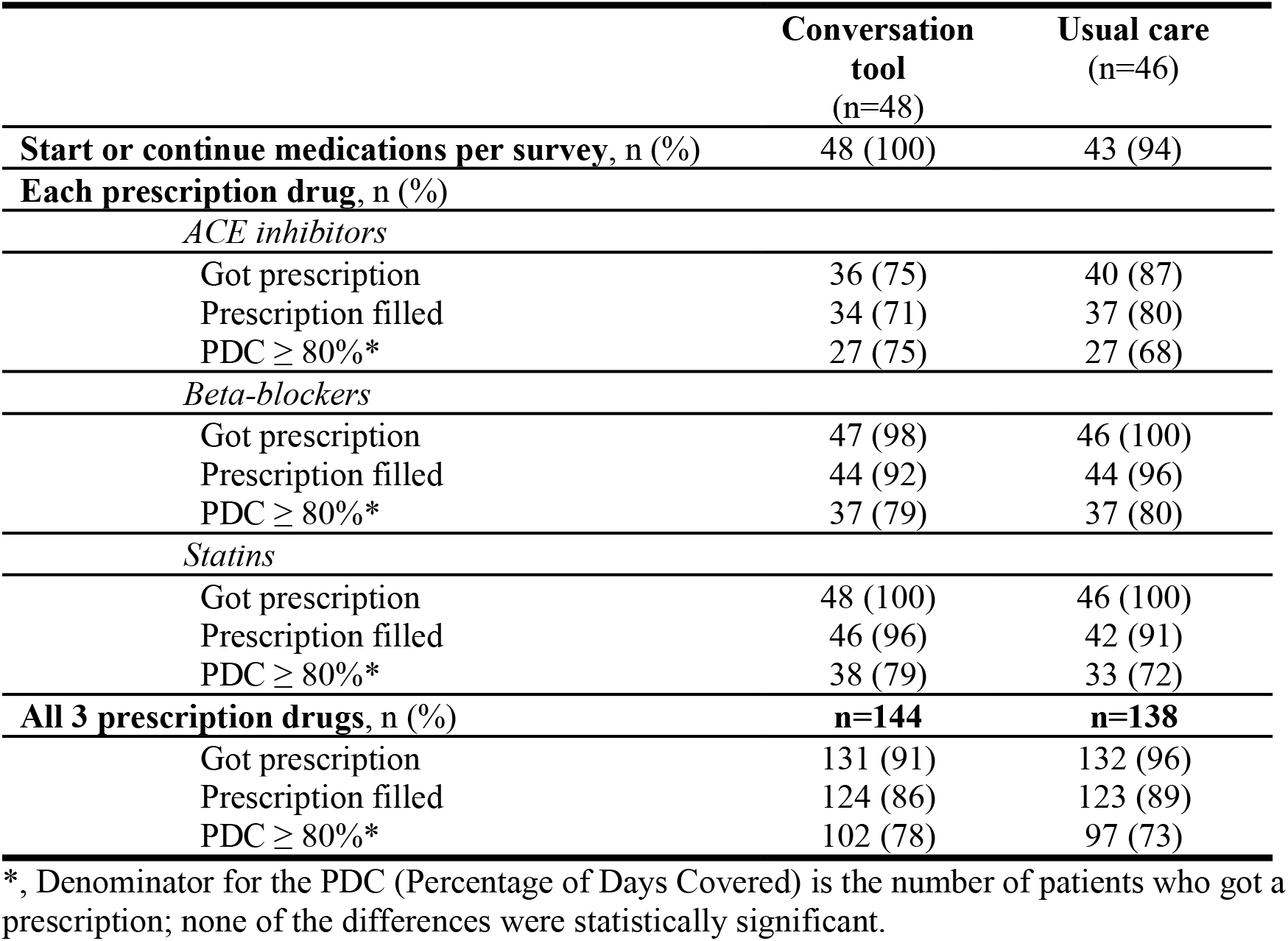
Post myocardial infarction adherence to medications

|  | **Conversation**<br>**tool**<br>**(n=48)** | **Usual care**<br>**(n=46)** |
| --- | --- | --- |
| **Start or continue medications per survey, n (%)** | 48 (100) | 43 (94) |
| **Each prescription drug, n (%)** |  |  |
| *ACE inhibitors* |  |  |
| Got prescription | 36 (75) | 40 (87) |
| Prescription filled | 34 (71) | 37 (80) |
| PDC ≥ 80%* | 27 (75) | 27 (68) |
| *Beta-blockers* |  |  |
| Got prescription | 47 (98) | 46 (100) |
| Prescription filled | 44 (92) | 44 (96) |
| PDC ≥ 80%* | 37 (79) | 37 (80) |
| *Statins* |  |  |
| Got prescription | 48 (100) | 46 (100) |
| Prescription filled | 46 (96) | 42 (91) |
| PDC ≥ 80%* | 38 (79) | 33 (72) |
| **All 3 prescription drugs, n (%)** | **n=144** | **n=138** |
| Got prescription | 131 (91) | 132 (96) |
| Prescription filled | 124 (86) | 123 (89) |
| PDC ≥ 80%* | 102 (78) | 97 (73) |

\*, Denominator for the PDC (Percentage of Days Covered) is the number of patients who got a prescription; none of the differences were statistically significant.

Of the 106 patients, 6-month pharmacy records were obtained for 94 (thus 12 were lost to follow-up, 5 in the intervention and 7 in the usual care group). Overall, 86% of the prescriptions (124 out of 144) were ever filled for the *AMI Choice* group and 89% of the prescriptions (123 out of 138) were ever filled in the usual care group. The percentage of patients fully adherent to their medications (PDC ≥ 80%) was similar (78% in the conversation tool group vs. 73% in the usual care group; **Table 4**). At 6 months, aspirin use was similar in both groups (conversation tool, 48/50 (96%); usual care, 45/49 (92%)).

## DISCUSSION

In this pilot randomized trial, use of the *AMI Choice* conversation tool in the hospital setting by nurse practitioners and registered clinical nurses caring for the patient proved feasible and effective in transferring key knowledge, improving decisional comfort, and enabling greater patient involvement in shared decision making.

Low treatment fidelity to life-saving medication regimens after AMI at 1 year may be due to structural factors (access to and cost of medications) and volitional factors (treatment is not consistent with patients’ knowledge and preferences). Choudhry and colleagues evaluated the impact of eliminating copayments for medications for patients after AMI in whom adherence to medications ranged from 36% to 49%.^7^ In this trial, the intervention led to modest (≤ 7%) improvements in adherence. A more recent trial focused on dual antiplatelet therapy found a similarly favorable result (i.e., improvements in persistence of about 10%, to about 54-59%) with copayment assistance vouchers.^27^ Beyond addressing structural factors, SDM may address volitional factors by improving patient knowledge of the benefits and risks of taking medications and by improving patient involvement in treatment decisions. A recent small, randomized trial found a small but significant beneficial effect of a video-based education program for patients post AMI on self-reported adherence at 3 months.^28^ In our trial, adherence to medications at 6-months, while inadequate, was relatively high, ranging from 68% to 80%, and was comparable between the conversation tool and usual care groups, leaving little room for improvement.

However, in a more recent pragmatic trial in a cohort of patients in which only 36% exhibited full adherence to medications post AMI, mailed reminders and phone calls were also ineffective.^29^ Conversely, in a randomized trial enrolling older patients with low adherence and comparing usual care with and without a nursing intervention (assessment of problems with adherence and targeted education) at 3 months post AMI found that the intervention almost doubled adherence (measured through self-report and pharmacy records) from 22% to 52%.^30^ To our knowledge, the trial reported herein is the only one deploying an SDM intervention to improve adherence in the post AMI period, joining a body of evidence that offers an inconsistent effect of SDM on adherence and outcomes.^31-35^ Future studies that aim to improve fidelity to post AMI treatments may need to enroll patients at highest risk of nonadherence and design, implement, and evaluate interventions that address both structural and volitional factors while addressing the practical challenges of making treatment programs fit in daily life routines.^36^

### Limitations

The study was conducted by personnel who participated in the development of the conversation tool and among clinicians practicing in an academic medical center. Furthermore, the study enrolled patients with limited socioeconomic diversity and high adherence to medicines. All these affect the applicability of this pilot trial to other settings and populations. This trial was randomized at the patient-level. However, the risk for contamination was mitigated since the clinicians delivering the intervention could not generate a tailored conversation tool for patients randomized to usual care. The conversation tool was used once during the hospitalization and was not used again during follow-up to support ongoing decision-making. Keeping these limitations in mind, the results suggest *AMI Choice* is a feasible and effective intervention to promote shared decision making among AMI survivors. Further research to establish its effect on other measures of quality beyond patient-centered care is warranted.

## CONCLUSIONS

Our conversation tool, *AMI Choice*, was effectively used by nurse practitioners and registered clinical nurses caring for patients hospitalized following AMI. Patients randomly assigned to the *AMI Choice* conversation tool gained greater knowledge than usual care patients, as their clinicians made substantial efforts to involve them in their medication treatment decisions.

Medication adherence at 6 months was comparable for both the conversation tool and usual care groups. Future SDM trials evaluating strategies to improve fidelity to post AMI regimes will need enroll participants at high risk for nonadherence and intervene not only at the time of discharge but also provide ongoing support for the implementation of the medication regimen over time.

This may include the repeated use of the *AMI Choice* conversation tool.

## Data Availability

All data produced in the present study are available upon reasonable request to the authors

